# Quantifying the impact of the COVID-19 pandemic on the Scottish accident and emergency landscape

**DOI:** 10.1101/2024.12.20.24319411

**Authors:** Hui Pheng Teoh, Kasia Banas, Christopher Aldous Oldnall

**Affiliations:** The Usher Institute, The University of Edinburgh, Edinburgh Bioquarter, 9 Little France Road, Edinburgh, EH16 4UX, UK; School of Mathematics, The University of Edinburgh, James Clerk Maxwell Building, Peter Guthrie Tait Rd, Edinburgh, EH9 3FD, UK

## Abstract

**Background:** The COVID-19 pandemic, declared in March 2020, is suspected to have greatly impacted Scotland’s accident and emergency (A&E) services. Stringent public health measures, including lockdowns, heightened pressures on A&E departments, but their long-term effects remain understudied. This study examines how the pandemic influenced A&E attendances over a broader time-frame.

**Methods:** Secondary data from Public Health Scotland (2018–2022) on monthly A&E attendances was analysed, standardised per 1,000 population using census data. Choropleth maps visualised A&E attendances and wait times across health boards during key pandemic periods. A Poisson generalised linear model (GLM) assessed the influence of COVID restrictions, demographics, and service factors on attendance rates.

**Results:** A&E attendances dropped from 2018 to 2022, with NHS Lanarkshire (-32.7%) and NHS Borders (-27.9%) seeing the largest declines. Despite reduced attendances, the percentage of patients seen within four hours also dropped (-29.4% in NHS Lanarkshire). The Poisson GLM confirmed that COVID-19 restrictions influenced attendance rates with a lower incident rate (IRR: 0.89, 95% CI: 0.88–0.90) during lockdowns and an increased incident rate (IRR: 1.04, 95% CI: 1.03–1.05) during easing periods. Health board-level effects varied substantially.

**Discussion:** COVID-19 disrupted A&E services, causing fluctuating attendances and worsening wait times. Large health board-level variations suggest local policies, behaviours, and existing pressures significantly influenced outcomes. These findings highlight the need to address systemic issues alongside localised strategies for future resilience.

## 1 Background

In March 2020, the World Health Organisation (WHO) declared a global coronavirus (COVID-19) pandemic [1]. In response, the United Kingdom (UK) and Scottish governments imposed stringent public health measures, including lockdowns, to mitigate the spread of the virus [2, 3]. These measures included the a redistribution of resources from a range of services, including A&E, towards COVID-designated wards. It is believed that these measures exacerbated existing challenges in the health service, particularly in accident and emergency (A&E) departments, which were already under significant pressure due to increasing demand over the preceding years [4–6]. Prolonged A&E waiting times are associated with poorer clinical outcomes, with estimates suggesting one avoidable patient death for every 72 patients who wait 8-12 hours from arrival [7].

The management of UK National Health Service (NHS) is devolved across the four nations—England, Scotland, Wales, and Northern Ireland—with healthcare devolvement to Scotland taking place in 1999, with full fiscal and public health devolution occurring in 2012, granting it greater autonomy over health service management [8]. While multiple studies have explored A&E activity in England [9–18], there remains a significant gap in research focusing on Scottish A&E data [19, 20]. Comparative analysis between the four UK nations is challenging due to variances in service delivery, data collection methodologies, and definitional standards [21, 22]. This study seeks to explore A&E trends in the unique context of Scotland’s healthcare with a focus on the impact of COVID-19 on Scottish A&E attendances.

In February 2024, the Office for National Statistics (ONS) published a report on A&E waiting times, which included a limited selection of Scottish data to compare A&E performances across the UK from 2013 to 2023 [23]. The most comprehensive public resource, to our knowledge, that focuses on Scottish A&E activity remains the interactive charts provided by Public Health Scotland (PHS) [22]. PHS plays a critical role in collating and disseminating health-related data and intelligence across Scotland, informing decisions that impact public health. The statistical practices of PHS are regulated by the Office for Statistics Regulation (OSR) to ensure accuracy and reliability [22, 24]. Both the ONS and PHS analyses have identified an upward trend over the past decade in the percentage of patients waiting longer than four hours in Scottish A&E departments [22, 23].

The impact of the COVID-19 pandemic on Scottish A&E activity has been examined in two key studies, both of which focused on the first 12 months of the pandemic [19, 20]. These studies compared A&E attendance numbers during periods of government-imposed restrictions with average attendance figures from the corresponding periods in 2018-2019. The findings indicated a significant reduction in A&E attendances during the periods of restriction. This trend was observed across various demographic groups, including sex, socioeconomic status, and most age categories, although children aged 0-14 years consistently exhibited notably lower A&E attendances throughout the study period [20]. These early findings highlight the need for further investigation into the long-term effects of the pandemic on Scottish A&E services, especially as the pandemic response evolved over time.

This is the first study to analyse the long-term impact of COVID-19 on Scottish A&E services over five years. We seek to develop a comprehensive understanding of the key trends and factors, for which data exists, influencing Scottish A&E attendances and waiting times. The analysis employs population estimates to compare A&E attendance rates and waiting times across different Scottish health boards, allowing for insights into regional variations and the effectiveness of local healthcare responses. The insights derived from this analysis will be instrumental in benchmarking Scottish health boards’ performance, optimising A&E services, and preparing for future healthcare challenges, such as a service impacting epidemic or pandemic.

## 2 Methods

This study utilised secondary data collected and published by Public Health Scotland (PHS). We obtained descriptive statistics for the data to gain a baseline understanding of Scottish A&E activity from 2018 to 2022. For this we define the A&E attendance rate per 1,000 population as,

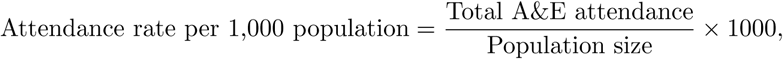

where the population size is matched to the relevant data per individual health boards utilising the 2022 census data [25] – enabling population scaling for each health board for the relevant years. The percentage of A&E patients seen within 4 hours by individual health boards is calculated in the following way,

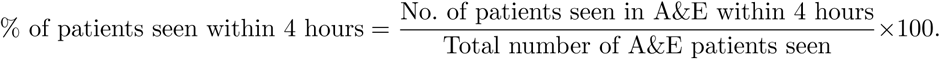

### 2.1 Data Source and Structure

The dataset utilised in this study encompasses monthly accident and emergency (A&E) statistics from 1 January 2018 to 31 December 2022, covering 30 large consultant-led emergency departments (EDs) and 64 minor injuries/other units (MIU/Other) across Scotland [26]. Throughout this article A&E refers to all emergency health departments, whilst ED and MIU/Other refer to the specific department types as aforementioned. These data points are aggregated to include all A&E sites in Scotland. Planned attendances following contact with Flow Navigation Centres, established as part of the Redesign of Urgent Care (RUC) program implemented in December 2020, were not part of the original dataset [22].

The planning, commissioning and delivery of healthcare services in Scotland are the responsibility of 14 regional NHS health boards that report to the Scottish government [27, 28]. The boundaries of the NHS boards are based on the combined area of each local authority that they serve [28]. The dataset was enriched by incorporating health board population estimates (per year) from the 2022 census data [25]. This allowed for standardisation, across time, of attendance rates per 1,000 population. Additionally a COVID-19 time-period indicator variable was introduced into the data to indicate four different states of COVID-19 related restrictions; ‘No (or) most restrictions removed’, ‘Easing of restrictions’, ‘Tightening of restrictions’, ‘Lockdown’. Figure 1 provides a timeline of the indicator values.

**Fig. 1:**
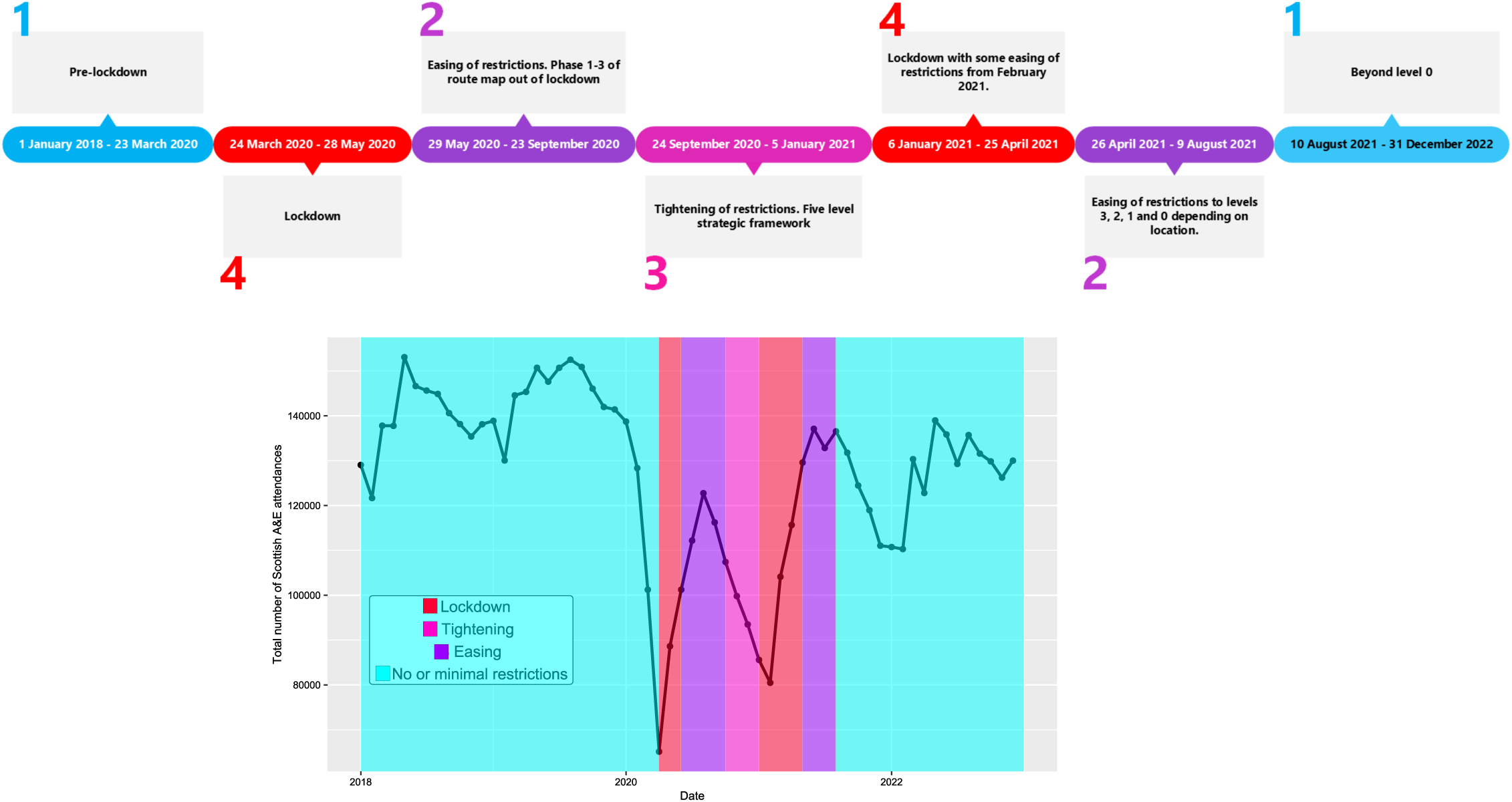
Attendances and COVID encoding. Levels 1 through 4 are used to represent: ‘No (or) most restrictions removed’, ‘Easing of restrictions’, ‘Tightening of restrictions’, ‘Lockdown’. These are detailed further in the timeline on the top row, with an overlay on the total number of attendances across Scotland on the bottom row.

The raw data were obtained in .csv and .xlsx formats [22, 26], and were processed using both R (version 4.3.1)/RStudio (2024.09.2 Build 375) and Python (version 3.9.15). The datasets included: the number of A&E attendances, percentage of patients seen within 4 hours, age, sex, health board, department type, arrival hour, day of the week, month, and deprivation status. Variables are detailed in a flow diagram and described in Appendix A.

Data cleaning involved the exclusion of entries with missing or ambiguous values. Quantitative descriptive statistics, including measures of central tendency (mean, median) and dispersion (standard deviation), were calculated to provide baseline insights into the dataset. The normality of the data distribution was assessed using visual inspection (Q-Q plots) and histograms, which indicated that the data did not follow a normal distribution, necessitating the use of non-normal methods in subsequent analyses. In addition the KS-test was employed, but its limitation to application of continuous data noted [29].

### 2.2 Choropleth

Choropleth maps provide an intuitive visualisation of spatial patterns by grouping and assigning attribute values a different colour, shade or pattern [30]. In this study, choropleth maps were utilised to illustrate differences in A&E activity across the Scottish NHS boards over time. Choropleth maps for 2018, 2020 and 2022 depicting A&E (ED and MIU/Other) attendance rates and the percentage of patients seen within 4 hours by health board were produced using the NHS Scotland 2019 health board shape files downloaded from the Scottish Government Spatial Data site [28]. 2018, 2020 and 2022 were chosen to represent the periods before, at the start and end of the COVID-19 pandemic.

### 2.3 Generalised Linear Modelling (GLM)

Our baseline knowledge from the choropleth and summary statistics is drawn together formally with linear regression to measure the impact that demographics (sex, age, deprivation status), health board, department type, temporal measurements (attendance by day of the week, month of the year and arrival hour) and COVID-19 factors had on A&E activity. Generalised linear models (GLMs) are a flexible generalisation of ordinary linear regression that allows for response variables to have error distribution models other than a normal distribution. This flexibility is particularly useful for modelling count data, such as A&E attendances, which are non-negative and discrete.

Given the nature of the data, a Poisson regression model, a type of GLM, was selected. Poisson regression is appropriate for modelling event count data, where the mean of the response variable is assumed to be equal to its variance. In situations with overdispersion—where the variance significantly exceeds the mean—alternative models, such as negative binomial regression, could typically be considered [31].

The Poisson regression used in this study assumes *Y_i_* ∼ Poisson(*µ_i_*), with E[*Y_i_*] = *µ_i_* and Var(*Y_i_*) = *µ_i_*. We take *µ_i_* to be the expected count of A&E attendances for the *i*-th observation. The model form is as follows,

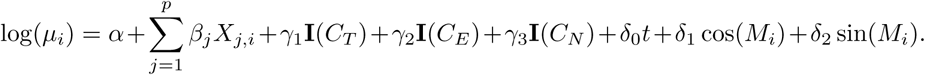

In the model, *α* represents the intercept, or a baseline log-expected count. 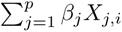 represents the sum of the effects of the continuous proportional predictor variables. Each predictor variable in this case represent proportions within different categories (sex, age group, deprivation status, day of the week, department type and health boards).

All *γ*-type variables represent the effects of different COVID periods as indicated on Figure 1 and expanded upon in Appendix B. The indicator functions **I**(·) are equal to 1 during the ‘lockdown’, ‘tightening’ and ‘easing’ periods respectively, and 0 otherwise, with ‘no restriction’ being the reference category for the categorical variable.

*δ*_0_*t* represents the linear effect of time (measured in days from the start date 01.01.2018), with *δ*_0_ indicating how the log-expected count changes with time. The terms *δ*_1_ cos(*M_i_*) + *δ*_2_ sin(*M_i_*) capture cyclic monthly seasonality, where *M_i_* represents the month of observation *i*. Together, these terms model the periodic fluctuation in counts across months, with coefficients *δ*_1_ and *δ*_2_ controlling the amplitude and phase of the seasonal cycle.

The model was built using data from January 2018 to December 2022, as detailed information on patient demographics and arrival times is only published from 2018. The chosen model incorporated sex, age, deprivation status, day of the week, department type, NHS health board, encoding for the different stages of lockdown, tightening and easing of restrictions and time. Time refers to the number of days from the start of the model (January 2018). The Akaike Information Criterion (AIC) and McFadden’s R-squared were used to select factors to include in the model, with lower AIC values and higher McFadden’s R-squared values indicating better model fit.

To evaluate the net effect of each of the categorical variables on the outcome over time, we compute the weighted multiplicative effect at each time point. This utilises the coefficients from the log-model and the proportions of each category (or the assigned indicator in the case of the COVID variable). Per categorical variable this is calculated as,

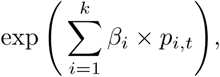

where *k* is the number of categories in the variable, *β_i_* is the estimated coefficient for the *i*-th category in the variable and *p_i,t_* is the proportion for the *i*-th category at the time point *t*. This provides a single, interpretable measure of the net effect of the variable, which can be interpreted as the expected multiplicative impact of a variable on the outcome. Producing this calculation across all time-points allows for dynamic evolution of the net effect of the variable, demonstrating the time-varying measure of its impact.

## 3 Results

### 3.1 Summary statistics find wide ranging attendances

There were 4,456 entries in the A&E monthly attendance and waiting times dataset from 1 January 2018 to 31 December 2022. Appendix C provides descriptive statistics (minimum, mean, median, maximum, standard deviation, and variance) for all A&E departments, as well as a breakdown by ED and MIU/Other. During this period, EDs recorded higher attendances but lower percentages of patients seen within four hours compared to MIU/Other.

The number of Scottish A&E attendances is discrete, non-continuous data and limited to non-negative values. A histogram and Q-Q plot of A&E attendances from 1 January 2018 to 31 December 2022 (Appendix C) indicate that the data do not follow a normal distribution. Although the K-S test returned a p-value of 0.08486 (*>* 0.05), suggesting normality, this was disregarded due to the test’s limitation of application to continuous distributions.

While the Poisson regression assumption of equal mean and variance was violated (*µ* = 1, 711; *σ* = 5, 116, 963), the decision was made to proceed with the modelling. This was based on the understanding that the variance, given the wide spread of the distribution, may not be a reliable indicator of model appropriateness in this context. It was determined that the Poisson model would still provide valuable insights, and overdispersion could be managed through robust standard errors.

### 3.2 Choropleths demonstrate unexpected trends in attendance rate and patients seen within 4 hours around COVID-19

Figure 2 provides a visual comparison of A&E performance across health boards in Scotland in 2018, 2020, and 2022, highlighting the trends in both attendance rates and the percentage of patients seen within four hours. The choropleth maps illustrate how these measures evolved over time, particularly during and after the COVID-19 pandemic.

**Fig. 2:**
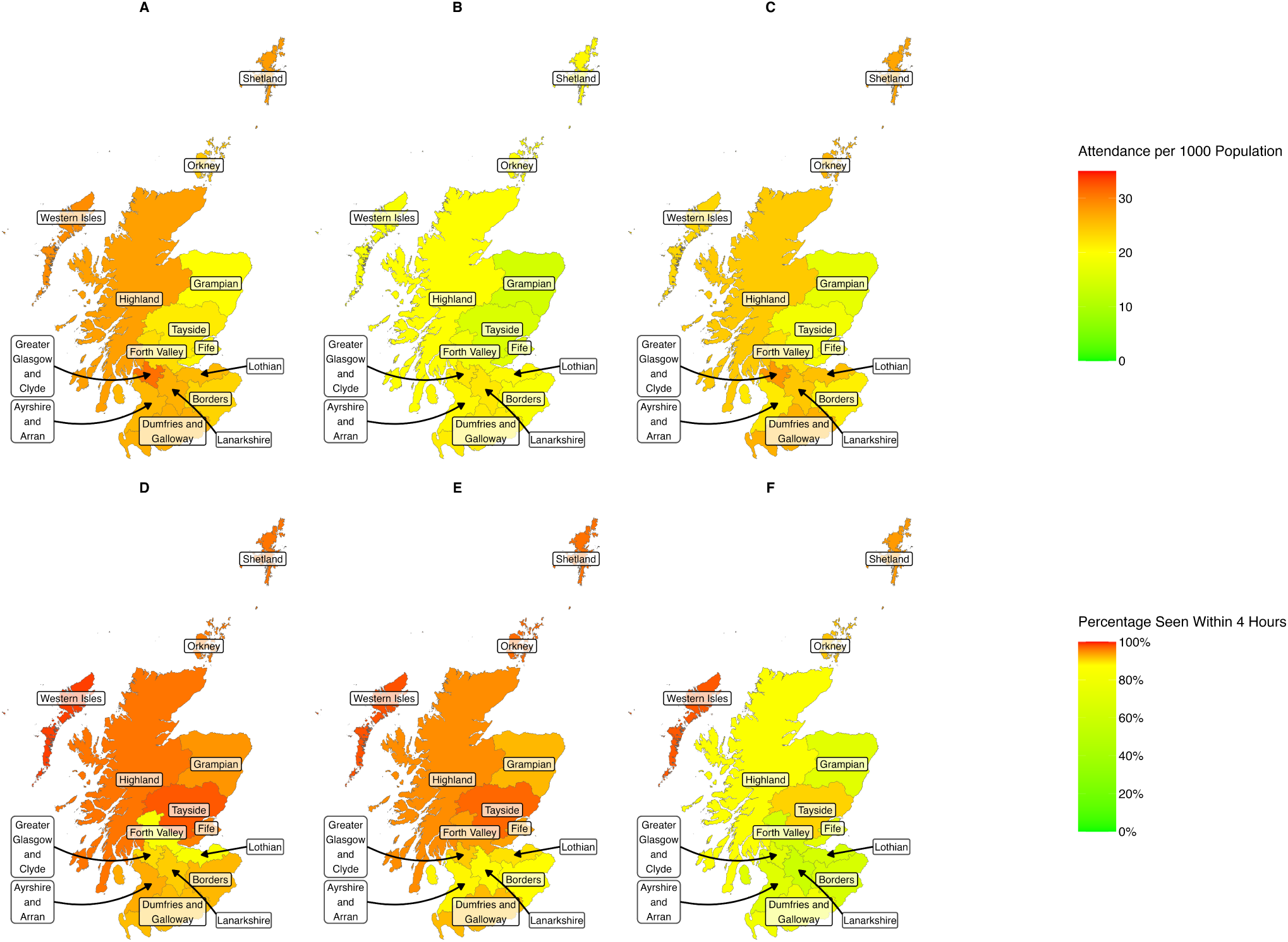
A&E attendance rates and waiting times in 2018, 2019 and 2020. Choropleth maps demonstrating trends in A&E attendance rates and percentage of patients seen within four hours across Scottish health boards in 2018 (panels A, D), 2020 (panels B, E), and 2022 (panels C, F). Panels A-C show attendance rates, while panels D-F show the percentage of patients seen within four hours.

The data show that attendance rates dropped across almost all health boards between 2018 and 2022, as indicated by the change in colours from yellow/orange to green across most regions. Orkney was the only health board to display a slight increase in attendance rates during this period. Despite the reduction in attendance, waiting times worsened, with the percentage of patients seen within four hours declining in all health boards, shown by the shift towards more yellow and green tones in 2022 compared to earlier years.

In 2020, at the height of the COVID-19 pandemic, there was a noticeable decline in attendance rates across all health boards (Figure 2, panel B). This is represented by the lighter yellow hues indicating lower attendance. However, by 2022, while attendance rates had increased relative to 2020, they still had not returned to 2018 levels, as reflected by the persistence of light orange and yellow colours in many areas (panel C).

The percentage of patients seen within four hours also exhibited significant variation between health boards over the same period. In 2020, some health boards—including NHS Forth Valley, NHS Greater Glasgow and Clyde, NHS Lothian, NHS Orkney, and NHS Shetland—improved their performance in terms of patients seen within four hours compared to 2018 (panel E), as indicated by the dark red/orange colours in these regions. In contrast, other boards, such as NHS Ayrshire and Arran and NHS Borders, saw a decrease in this metric during the same period, shown by the shift to yellow tones.

By 2022 (panel F), all health boards had experienced a drop in the percentage of patients seen within four hours compared to both 2018 and 2020. The most substantial reductions from 2020 to 2022 were observed in NHS Lanarkshire and NHS Forth Valley, where the percentage of patients seen within four hours dropped by 29.4% and 29.0%, respectively, as indicated by the lighter yellow/green tones in these regions. When comparing 2018 to 2022, NHS Lanarkshire experienced the largest overall decline, with a drop of 32.7%, followed by NHS Borders, which saw a 27.9% reduction.

These trends demonstrate that while attendance rates initially dropped during the COVID-19 pandemic, waiting times worsened across the board, with fewer patients being seen within four hours as time progressed. This is especially evident in the sharp contrast between 2020 and 2022, where the maps show a widespread shift towards green and yellow tones, reflecting the increasing pressures on A&E services across Scotland’s health boards.

### 3.3 Generalised linear modelling highlights impact of COVID-19 restrictions on attendances

The Poisson generalised linear model (GLM) analysis revealed that COVID-19 restrictions, along with other demographic and service-related factors, significantly influenced A&E attendances. The analysis excluded arrival hour to focus on broader factors impacting attendance patterns.

COVID-19 restrictions had a marked effect on attendance rates. During periods of easing of restrictions, the incident rate ratio (IRR) was 1.040 (95% CI: 1.029, 1.051). As restrictions tightened, the IRR decreased to 1.001 (95% CI: 0.989, 1.014). With the strictest lockdown restrictions, the IRR remained decreased down to 0.892 (95% CI: 0.880, 0.904). These results highlight the significant and lasting influence of public health measures on healthcare demand, with attendances fluctuating throughout COVID-19 periods.

For socio-demographic factors, males had a lower likelihood of attending A&E compared to females, with IRRs of 6.715 (95% CI: 4.885, 9.231) and 10.992 (95% CI: 7.914, 15.268) respectively. Age was also significant, with younger age groups attending at higher rates than most older individuals, the exception being the 75 plus group. Those under 18 had an IRR of 7.030 (95% CI: 6.114, 8.083), while the 18–24 age group showed an IRR of 6.262 (95% CI: 4.193, 9.353). Those in the 75 plus group were the at highest risk of attending with an IRR of 41.338 (95%: 20.389, 83.808). Deprivation, as measured by the Scottish Index of Multiple Deprivation (SIMD), revealed a gradient of attendance, with the least deprived group (SIMD 5) having the highest rate of attendance with an IRR of 218.153 (95% CI: 82.202, 578.950), with SIMD 2 and SIMD 1 also having an IRR over 1 (10.874 95% CI: (4.571, 25.870) and 2.511, 95% CI: (1.369, 4.605) respectively).

A&E service-related factors also influenced attendance patterns. Attendances varied across the week, with weekends experiencing lower rates. Tuesday and Saturday exhibited the lowest IRRs, at 0.867 (95% CI: 0.802, 0.938) and 1.176 (95% CI: 1.076, 1.287) respectively. Emergency departments had a lower IRR of 7.455 (95% CI: 5.919, 9.391) compared to minor injuries units, with an IRR of 29.238 (95% CI: 21.342, 40.054). Variations across NHS health boards were also notable with extreme large and small values observed in Orkney and Shetland, respectively. The time variable, while significant, showed only a marginal effect on attendances, with an IRR of 1.00 (95% CI: 1.00, 1.00). The negative coefficients of the monthly cosine and sine terms indicate a seasonal decline in attendance during specific months of the year.

Figure 3 demonstrates that the COVID-19 related factors have a larger (in magnitude) net effect through time than other factors included in the model. One exception to this is in the net effect of the health board variables, whereby we see a drop (0.762) and spike (1.283) in the health board variables contribution to the model during the earlier months of the COVID pandemic. There is also a drop visible on the net effect of the department type variables.

**Table 1:** Poisson GLM Results for A&E Attendances (Proportional Data)

| Predictor | Coefficient (log-scale) | IRR (Exp($\beta$)) | 95% CI on IRR |
| --- | --- | --- | --- |
| Intercept | 5.384 | 217.976 | (168.074, 282.695) |
| **COVID Period [Categorical]** |  |  |  |
| No (or) Most restrictions removed (ref) | — | 1.000 | — |
| Easing of restrictions | 0.039 | 1.040 | (1.029, 1.051) |
| Tightening of restrictions | 0.001 | 1.001 | (0.989, 1.014) |
| Lockdown | -0.114 | 0.892 | (0.880, 0.904) |
| **Sex** |  |  |  |
| Unknown Sex | 1.083 | 2.953 | (2.053, 4.247) |
| Male | 1.904 | 6.715 | (4.885, 9.231) |
| Female | 2.397 | 10.992 | (7.914, 15.268) |
| **Age Group** |  |  |  |
| Unknown Age | 1.083 | 2.953 | (2.053, 4.247) |
| Under 18 | 1.950 | 7.030 | (6.114, 8.083) |
| 18-24 | 1.835 | 6.262 | (4.193, 9.353) |
| 25-39 | -4.517 | 0.011 | (0.007, 0.018) |
| 40-64 | 1.955 | 7.066 | (4.804, 10.392) |
| 65-74 | -0.643 | 0.526 | (0.211, 1.306) |
| 75 Plus | 3.722 | 41.338 | (20.389, 83.808) |
| **Deprivation Status** |  |  |  |
| Unknown SIMD | 6.140 | 463.964 | (191.401, 1124.669) |
| SIMD 1 (most deprived) | 0.920 | 2.511 | (1.369, 4.605) |
| SIMD 2 | 2.386 | 10.874 | (4.571, 25.870) |
| SIMD 3 | -2.264 | 0.104 | (0.045, 0.240) |
| SIMD 4 | -7.183 | $7.592 \times 10^{-04}$ | $(2.888 \times 10^{-4}, 2.000 \times 10^{-3})$ |
| SIMD 5 (least deprived) | 5.385 | 218.153 | (82.202, 578.950) |
| **Day of the Week** |  |  |  |
| Monday | 1.773 | 5.890 | (5.463, 6.351) |
| Tuesday | -0.142 | 0.867 | (0.802, 0.938) |
| Wednesday | 1.365 | 3.915 | (3.567, 4.296) |
| Thursday | 0.855 | 2.350 | (2.130, 2.593) |
| Friday | 0.842 | 2.322 | (2.138, 2.521) |
| Saturday | 0.162 | 1.176 | (1.076, 1.287) |
| Sunday | 0.529 | 1.698 | (1.522, 1.894) |
| **Department Type** |  |  |  |
| Minor injuries unit | 3.375 | 29.238 | (21.342, 40.054) |
| Emergency department | 2.009 | 7.455 | (5.919, 9.391) |
| **NHS Health Boards** |  |  |  |
| Ayrshire and Arran (S08000015) | 6.935 | 1027.522 | (380.941, 2771.565) |
| Borders (S08000016) | -16.488 | $6.905 \times 10^{-8}$ | $(8.323 \times 10^{-9}, 5.729 \times 10^{-7})$ |
| Dumfries and Galloway (S08000017) | -8.110 | $3.004 \times 10^{-4}$ | $(7.652 \times 10^{-5}, 1.000 \times 10^{-3})$ |
| Forth Valley (S08000019) | 9.921 | $2.034 \times 10^4$ | $(7378.179, 5.609 \times 10^4)$ |
| Grampian (S08000020) | -9.998 | $4.551 \times 10^{-5}$ | $(1.844 \times 10^{-5}, 1.124 \times 10^{-4})$ |
| Highland (S08000022) | -0.931 | 0.394 | (0.140, 1.106) |
| Lothian (S08000024) | -8.540 | $1.955 \times 10^{-4}$ | $(1.021 \times 10^{-4}, 3.744 \times 10^{-4})$ |
| Orkney (S08000025) | 50.545 | $8.937 \times 10^{21}$ | $(3.048 \times 10^{19}, 2.621 \times 10^{24})$ |
| Shetland (S08000026) | -29.525 | $1.505 \times 10^{-13}$ | $(1.301 \times 10^{-15}, 1.741 \times 10^{-11})$ |
| Western Isles (S08000028) | 3.149 | 23.306 | (0.388, 1399.032) |
| Fife (S08000029) | -4.233 | 0.015 | (0.005, 0.046) |
| Tayside (S08000030) | 5.209 | 182.938 | (63.009, 531.130) |
| Greater Glasgow and Clyde (S08000031) | 5.936 | 378.447 | (211.410, 677.463) |
| Lanarkshire (S08000032) | 1.515 | 4.551 | (1.987, 10.425) |
| **Time** |  |  |  |
| Time | $3.327 \times 10^{-5}$ | 1.000 | (1.000, 1.000) |
| Cosine (monthly) | -0.021 | 0.980 | (0.974, 0.985) |
| Sine (monthly) | -0.019 | 0.981 | (0.978, 0.984) |
| **Model Fit Statistics** |  |  |  |
| AIC |  | 7220.731 |  |
| BIC |  | 6244.736 |  |
| McFadden |  | 0.957 |  |
| Log-Likelihood |  | -3571.366 (df=38) |  |

**Fig. 3:**
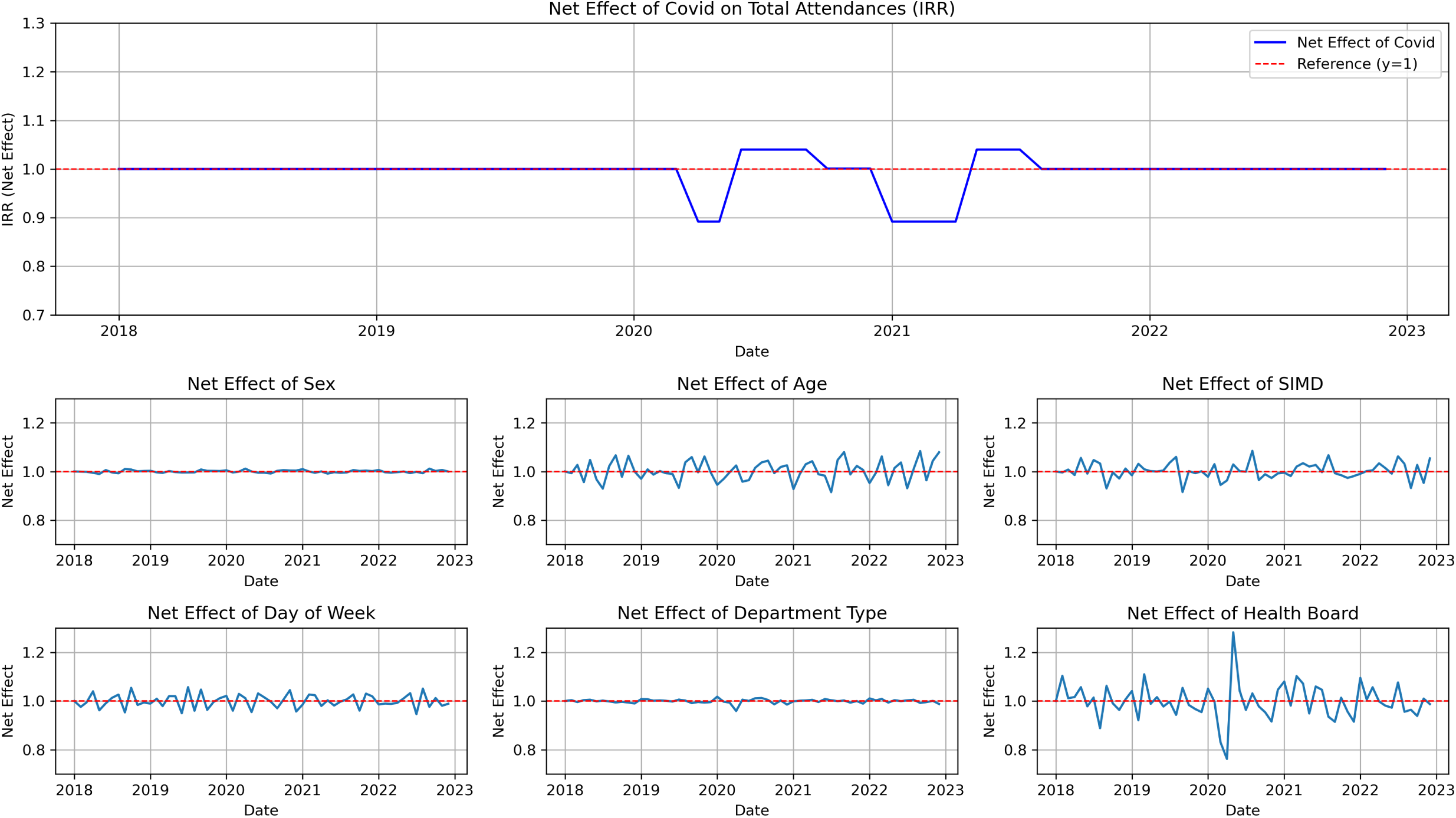
Net effects of variables on attendances across time. Here we see that there are varying stochastic net effects of the variables across time on attendances. Noticeably there is a sharp drop and spike in the health board net effect around the time of the COVID pandemic.

## 4 Discussion

### 4.1 Main Findings

This study analysed the impact of the COVID-19 pandemic on Scottish A&E services, revealing significant shifts in attendance patterns and service performance. A major finding is the sharp decline in the percentage of patients seen within four hours between 2018 and 2022, attributed to treatment backlogs and staff absences caused by illness or burnout. Urban health boards, such as NHS Lothian and NHS Greater Glasgow and Clyde, improved their performance, likely due to centralised resource allocation and planning. In contrast, rural boards, including Ayrshire and Arran and Lanarkshire, saw performance declines, reflecting limited resources and flexibility to adapt to demand surges.

GLM analysis showed that periods of tightening or easing of COVID-19 restrictions were associated with increased A&E attendances. While counter-intuitive, this rise may reflect a reduction in acute COVID-19 cases requiring emergency care, with the population returning to pre-pandemic behaviours alongside seasonal effects such as winter and festive periods. Public behaviour, influenced by perceived risks, likely played a role, sometimes preceding official restrictions. Demographic trends revealed that individuals aged 75 and older (IRR 41.338) had the highest attendance rates, consistent with their greater vulnerability to severe outcomes during the pandemic. These findings underscore how the pandemic strained A&E services and exposed disparities between urban and rural areas. They also highlight demographic vulnerabilities and the complex interplay between restrictions, seasonal factors, and public behaviour.

### 4.2 What is already known?

The COVID-19 pandemic placed unprecedented pressure on healthcare systems worldwide, exacerbating issues like treatment backlogs and staff shortages [5]. Existing literature identifies older populations as particularly vulnerable to severe health outcomes, contributing to higher morbidity and mortality rates during the pandemic [32].

### 4.3 What this study adds

This study reveals significant disparities between urban and rural A&E performance during the pandemic, highlighting the critical role of resource distribution and infrastructure. Urban centres benefitted from centralised resource allocation, while rural boards struggled with workforce shortages and inflexibility. The study also provides insights into how seasonal factors and public behavioural shifts intersected with policy measures, complicating direct attributions to restrictions alone. It further reinforces existing evidence of demographic disparities, showing that females and older adults were disproportionately represented in A&E attendances. These findings emphasise the need for tailored interventions that address local resource limitations and demographic vulnerabilities to improve service resilience during public health crises.

### 4.4 Limitations

This study’s reliance on monthly data limits its ability to capture short-term effects of policy changes. Behavioural shifts, driven by risk perception or media coverage, may have occurred before or after official lockdown periods, complicating causal attributions. Additionally, the exclusion of arrival hour from the GLM impacted model fit, potentially overlooking temporal dynamics such as time-of-day variations in attendance patterns. Seasonal factors, workforce issues, and local conditions—such as weather, flu seasons, and school terms—were not fully accounted for, introducing potential variability beyond the effects of COVID-19. Discrepancies between A&E activity and demographic datasets may have also introduced minor biases, though unlikely to affect key findings.

Future research should use more granular temporal data, such as weekly or daily figures, to better understand short-term impacts of restrictions and external factors. Incorporating qualitative insights from healthcare staff and patients could provide a richer understanding of the pressures on A&E services, complementing quantitative trends. This approach would allow for more effective interventions to address vulnerabilities exposed during the pandemic.

## Data Availability

All original data utilised in the study are available from the Scottish open data platform: https://www.opendata.nhs.scot/is/dataset/monthly-accident-and-emergency-activity-and-waiting-times
Proportional data created for the purpose of modelling is included as supplementary table 1 with the manuscript. Additionally it can be accessed at the GitHub repo: https://github.com/UoE-Public-Health-Modelling/AECOVID19

https://github.com/UoE-Public-Health-Modelling/AECOVID19

## 5 Ethics

All data utilised in this study are publicly available, and no identifiable information was included. This research builds upon a MSc dissertation, which received ethical approval from the Usher Masters’ Research Ethics Group (UMREG, ID: UM24237).

## 6 Acknowledgements

The authors would like to thank NHS Scotland and Public Health Scotland for their dedication to improving health outcomes and for making the data publicly accessible. Their efforts in data collection and transparency have been invaluable to the completion of this study.

## A Data Overview

Table 2 summarises the categorical variables used in the analysis, including their respective categories. Age is stratified into six groups, ranging from under 18 to 75 years and over. Arrival hour captures the time of presentation in 1-hour intervals over a 24-hour period, while day of the week specifies the distribution across seven days. Department type distinguishes between Emergency Departments and Minor Injuries Units or other services. Deprivation status is based on the quintiles of the Scottish Index of Multiple Deprivation, providing a measure of socio-economic status. Geographical variation is accounted for using Scottish NHS health boards, covering all 14 regions. Lastly, sex is classified as female or male. This categorisation enables a comprehensive exploration of trends and patterns across key demographic and operational factors. Only variables utilised in the study are listed in the table.

**Table 2:** List of categorical variables and their values. Only data sources utilised in this study are listed and detailed in the table.

| Variable | Values |
| --- | --- |
| Age | Under 18, 18-24, 25-39, 40-64, 65-74 years, 75 plus |
| Arrival hour | Arrival times based on 1-hour intervals throughout a 24-hour period |
| Day of the week | Monday, Tuesday, Wednesday, Thursday, Friday, Saturday, Sunday |
| Department type | Emergency department or Minor Injuries Unit/Other |
| Deprivation status | Scottish Index of Multiple Deprivation quintiles 1-5 |
| Scottish NHS health boards | Ayrshire and Arran, Borders, Dumfries and Galloway, Fife, Forth Valley, Grampian, Greater Glasgow and Clyde, Highland, Lanarkshire, Lothian, Orkney, Shetland, Tayside, Western Isles |
| Sex | Female, Male |

Figure 4 illustrates a data flow diagram that maps out the structure and connections between datasets related to A&E activity and associated demographics. At the centre is Public Health Scotland, serving as the primary source for A&E activity data and population estimates. On this central point, various datasets converge, providing detailed data on monthly and weekly A&E activity and waiting times, including measures such as attendance numbers, percentages over specific waiting time thresholds, and treatment location. Additional datasets focus on demographics (e.g., age, sex, deprivation status), referral sources, discharge information, and multiple attendances. The diagram also highlights Health Board population estimates, which provide detailed age group breakdowns. Arrows represent the flow of non-confidential data into the teams repository (hosted on GitHub).

**Fig. 4:**
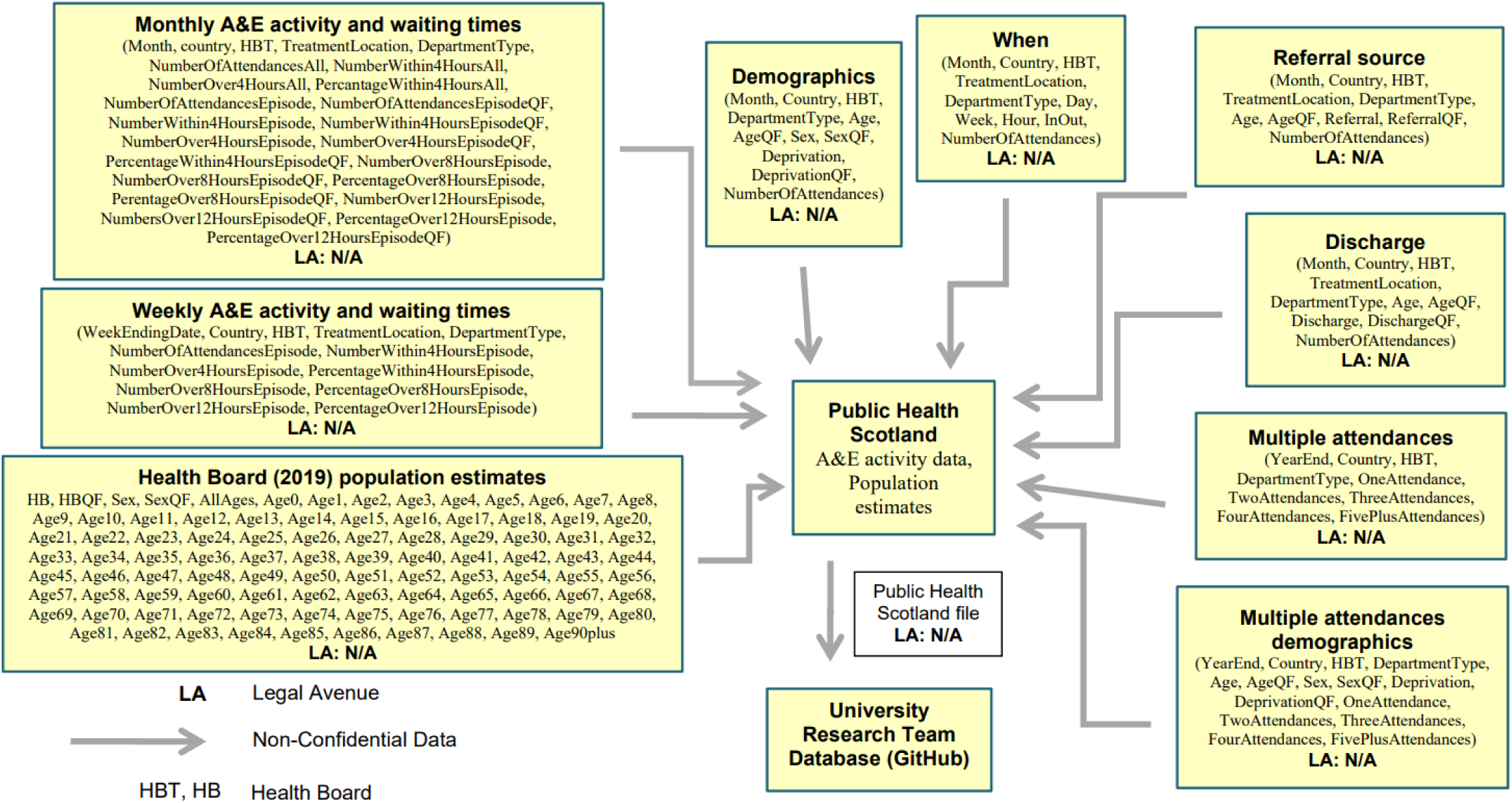
Data flow diagram. Flow of data sources used by Public Health Scotland to generate A&E data and its flow into our repository. All data sources are recorded in the diagram, but not all utilised in the study.

## B GLM Pandemic Date Encoding

**Table 3:**
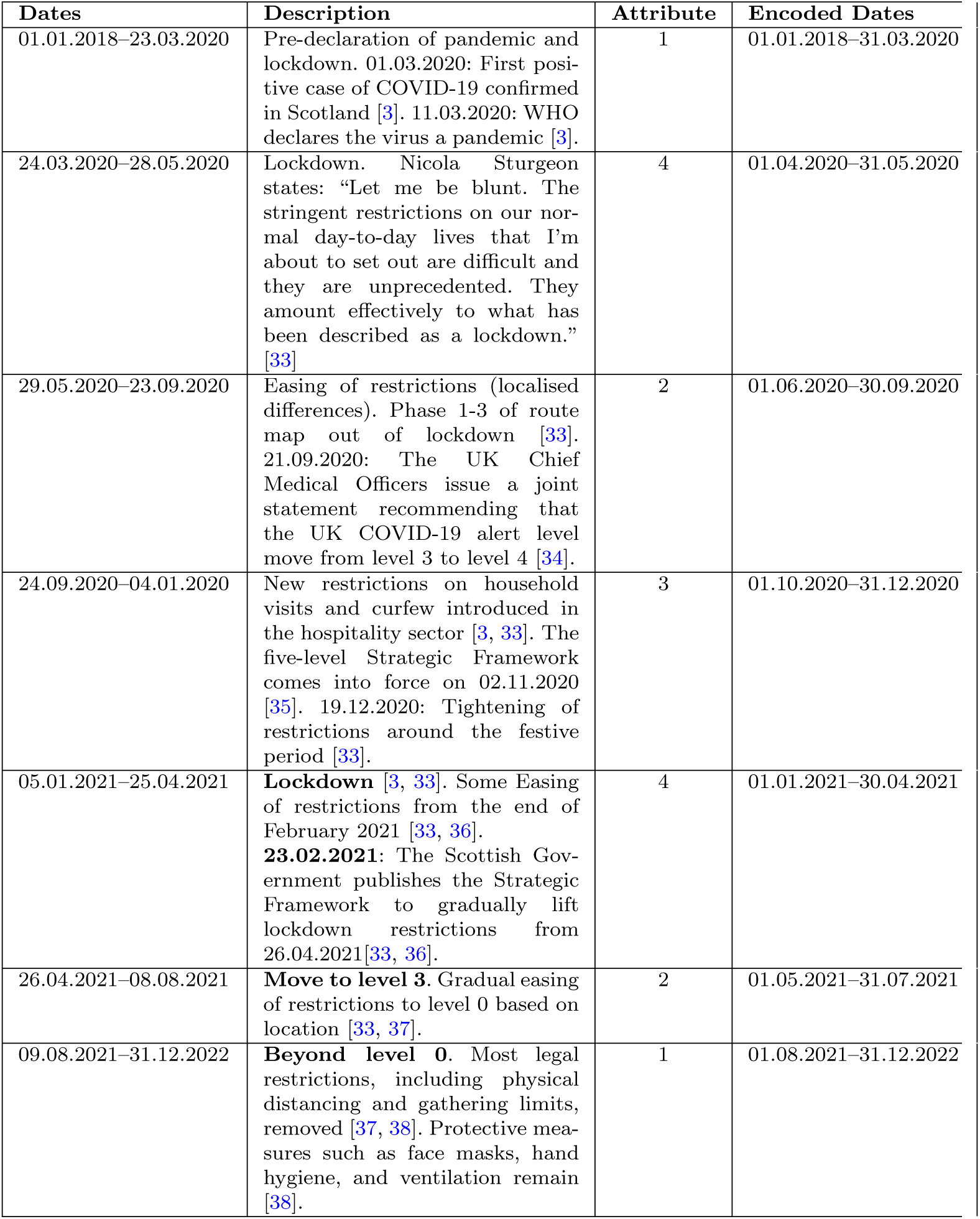
Timeline of COVID-19 restrictions. . Listed are a description of restriction conditions, attributes in the GLM, and encoded dates in the model.

| Dates | Description | Attribute | Encoded Dates |
| --- | --- | --- | --- |
| 01.01.2018–23.03.2020 | Pre-declaration of pandemic and lockdown. 01.03.2020: First positive case of COVID-19 confirmed in Scotland [3]. 11.03.2020: WHO declares the virus a pandemic [3]. | 1 | 01.01.2018–31.03.2020 |
| 24.03.2020–28.05.2020 | Lockdown. Nicola Sturgeon states: “Let me be blunt. The stringent restrictions on our normal day-to-day lives that I’m about to set out are difficult and they are unprecedented. They amount effectively to what has been described as a lockdown.” [33] | 4 | 01.04.2020–31.05.2020 |
| 29.05.2020–23.09.2020 | Easing of restrictions (localised differences). Phase 1-3 of route map out of lockdown [33]. 21.09.2020: The UK Chief Medical Officers issue a joint statement recommending that the UK COVID-19 alert level move from level 3 to level 4 [34]. | 2 | 01.06.2020–30.09.2020 |
| 24.09.2020–04.01.2021 | New restrictions on household visits and curfew introduced in the hospitality sector [3, 33]. The five-level Strategic Framework comes into force on 02.11.2020 [35]. 19.12.2020: Tightening of restrictions around the festive period [33]. | 3 | 01.10.2020–31.12.2020 |
| 05.01.2021–25.04.2021 | **Lockdown** [3, 33]. Some Easing of restrictions from the end of February 2021 [33, 36]. **23.02.2021**: The Scottish Government publishes the Strategic Framework to gradually lift lockdown restrictions from 26.04.2021 [33, 36]. | 4 | 01.01.2021–30.04.2021 |
| 26.04.2021–08.08.2021 | **Move to level 3**. Gradual easing of restrictions to level 0 based on location [33, 37]. | 2 | 01.05.2021–31.07.2021 |
| 09.08.2021–31.12.2022 | **Beyond level 0**. Most legal restrictions, including physical distancing and gathering limits, removed [37, 38]. Protective measures such as face masks, hand hygiene, and ventilation remain [38]. | 1 | 01.08.2021–31.12.2022 |

## C Summary Statistics

The descriptive statistics for A&E attendances and the percentage of patients seen within 4 hours from 1 January 2018 to 31 December 2022 are summarised in Table 1. The data is split across three categories: A&E overall, Emergency Departments (ED), and Minor Injuries Units (MIU)/Other, highlighting differences in minimum, median, mean, and variability of attendances as well as performance against the 4-hour waiting time standard. Figures 5 and 6 further explore the distribution of A&E attendances over this period. The histogram (Figure 5) shows the frequency of attendances, providing an overview of their distribution, while the Q-Q plot (Figure 6) assesses the normality of the data. Together, these visualisations and statistics give a comprehensive view of A&E performance and attendance trends over the study period.

**Table 4:** Descriptive statistics for A&E attendances and the percentage of patients seen within 4 hours. Statistics reported for A&E overall, Emergency Departments (ED), and Minor Injuries Units (MIU)/Other, for the period 1 January 2018 to 31 December 2022. The table includes measures of central tendency (mean and median), variability (standard deviation and variance), and the minimum and maximum values for attendances and 4-hour performance.

| **Attendances in a particular month** | **A&E** | **ED** | **MIU/Other** |
| --- | --- | --- | --- |
| Minimum | 1 | 195 | 1 |
| Median | 537 | 3,200 | 155 |
| Mean | 1,711 | 3,581 | 443.7 |
| Maximum | 11,579 | 11,579 | 4,394 |
| Standard deviation | 2,262 | 2,457 | 720 |
| Variance | 5,116,963 | 6,035,261 | 518,822 |
| **Percentage patients seen within 4 hours** | **A&E** | **ED** | **MIU/Other** |
| Minimum | 40.20% | 40.20% | 49.60% |
| Mean | 93.97% | 86.65% | 98.92% |
| Median | 98.90% | 90.70% | 100.00% |
| Maximum | 100.00% | 99.60% | 100.00% |

**Fig. 5:**
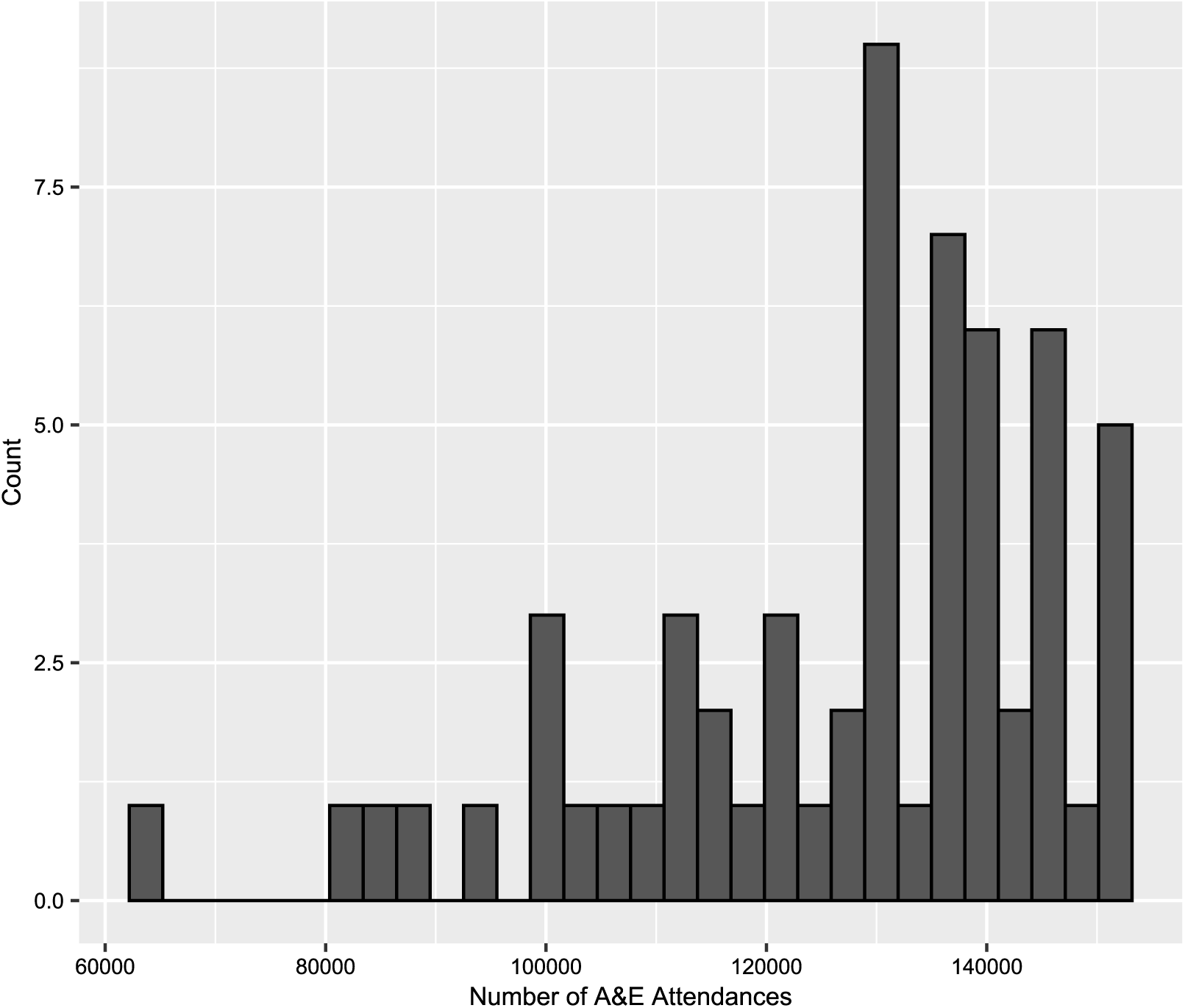
Histogram of A&E attendances from 1 January 2018 to 31 December 2022. The histogram highlights the frequency of lower and higher attendance levels, revealing the overall left-skewness and variability of the data.

**Fig. 6:**
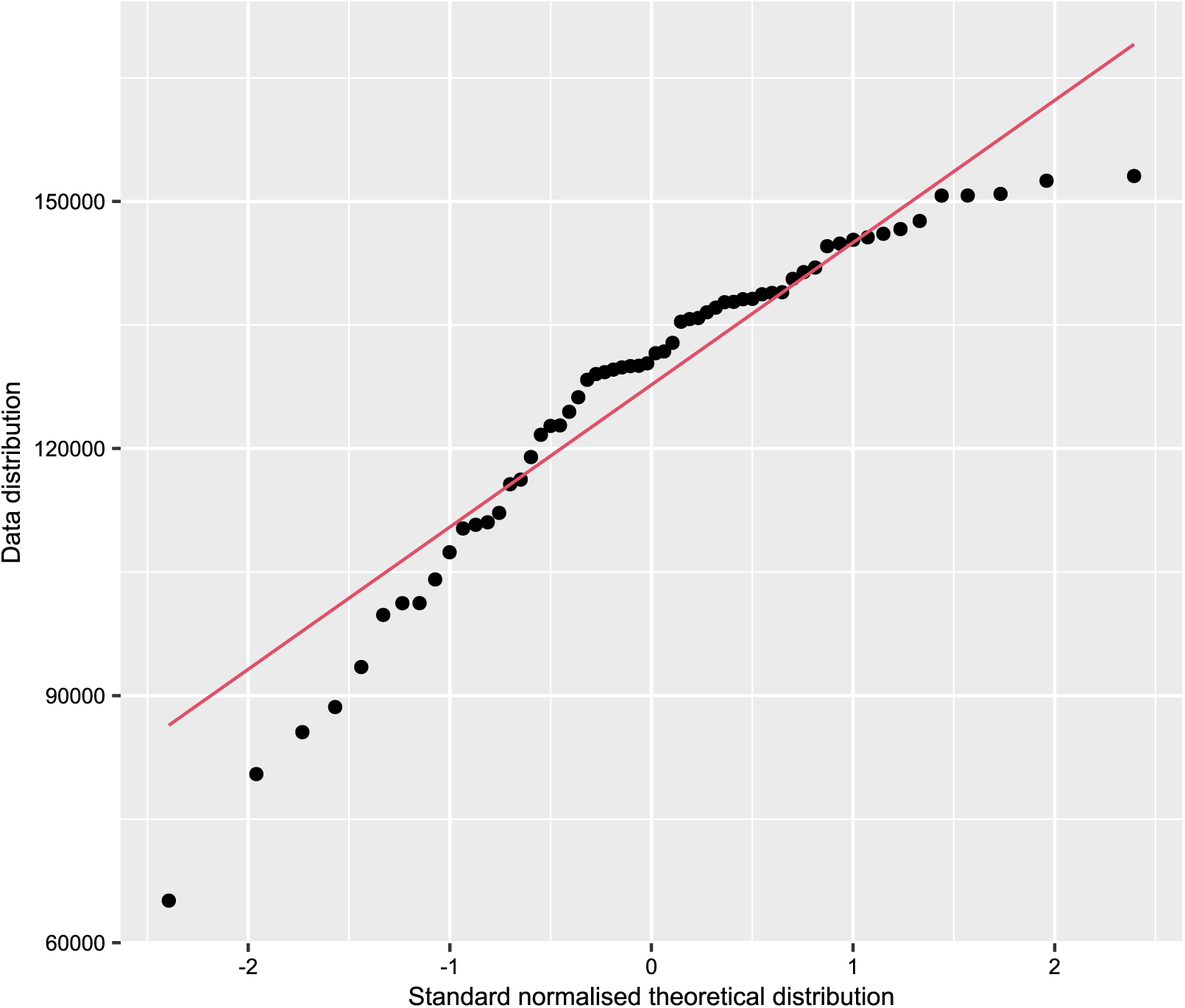
Q-Q plot of A&E attendances from 1 January 2018 to 31 December 2022. Plot illustrates the distribution of the observed data against a theoretical normal distribution. Deviations from the diagonal line indicate departures from normality, particularly in the tails of the distribution.

## D Supplementary Materials

### Supplementary Table 1: Proportional Data

Supplementary table 1 is the final data used to form the GLM. This data has 60 records (one per month) from the start of 2018 through to the end of 2022. There are then 43 columns, each of which relate to one variable used in the GLM – with the exception of the COVID period variable which has a singular column with categorical variables relating to each period.

